# Machine learning based time series forecasting of chemical levels from the East Palestine, Ohio train derailment site

**DOI:** 10.1101/2023.03.07.23286923

**Authors:** Vikas Ramachandra, Mohit Sethi

## Abstract

The paper presents a time series analysis of the air, water and soil data from the East Palestine, Ohio, train derailment site. The goal of the analysis is to investigate the temporal pattern of chemical pollutant levels in air, water and soil and to build machine learning and statistical models for forecasting various chemical concentrations over time.

## Introduction

The release of hazardous materials into the environment due to industrial accidents, transportation mishaps, and natural disasters poses a significant risk to public health and safety. Such incidents require prompt and effective response to mitigate the impact on the affected population and the surrounding environment. In the case of the train derailment that occurred in East Palestine, Ohio on 3 February 2023, 38 rail cars carrying hazardous material derailed, resulting in the release of hazardous material into the air, water, and soil.

The United States Environmental Protection Agency (EPA) and other agencies conducted air sampling in the area to monitor the concentration of the hazardous material and assess the potential risk to human health. The air sampling data collected at various locations in the vicinity of the incident site is a valuable source of information for evaluating the extent of contamination and predicting the future trend of the pollutant concentration. Time series analysis of the air sampling data can provide insights into the temporal behavior of the pollutant concentration and help identify any underlying patterns or trends.

In this paper, we perform a time series analysis of the air sampling data collected by the EPA following the East Palestine train derailment. Our objective is to identify any temporal patterns or trends in the pollutant concentration and evaluate the effectiveness of the remediation efforts. We focus on the “Result_Final_Txt” variable, which represents the concentration of the hazardous material in the air at different sampling locations over time. We then present our methodology for time series analysis and discuss the results of our analysis. Finally, we conclude with a discussion of the implications of our findings and the potential for future research in this area.

## Dataset

### EPA Lab Results: Air (through 02/22/23) - East Palestine, OH Response (csv) ^[1]^

(https://www.epa.gov/oh/air-sampling-data-east-palestine-ohio-train-derailment)

The dataset contains 1553 records starting from 04/02/2023 to 22/02/2023. The dataset contains following features: [‘Location’, ‘Samp_No’, ‘SampleDate_txt’, ‘SampleTime’, ‘Matrix’, ‘SampleMedia’, ‘Activity’, Analytical_Method’, ‘CAS_NO’, ‘Analyte’, ‘Result_Units’, ‘Reporting_Limit’, ‘Validation_Level’, ‘Result_Final_Txt’, ‘Result_Qualifier_Final’, ‘RL_Comparison’].

Our analysis is focussed on the ‘Results_Final_Txt’ which represents the concentration level of the hazardous chemical. The ‘Analyte’ and ‘CAS_NO” (CAS Registry Number) ^[4]^ help identify the chemicals. There are 49 unique chemicals. The records before 17/02/2023 are used for the training set(1036) and the remaining are used for the validation set(517).

### EPA Lab Results: Water (through 02/14/23) - East Palestine, OH Response (csv) ^[2]^

(https://www.epa.gov/oh/water-sampling-data-east-palestine-ohio-train-derailment)

The dataset contains 1822 records starting from 04/02/2023 to 17/02/2023. The dataset contains following features: [‘Location’, ‘Samp_No’, ‘SampleDate_txt’, ‘SampleTime’, ‘Matrix’, ‘SampleMedia’, ‘Activity’, Analytical_Method’, ‘CAS_NO’, ‘Analyte’, ‘Result_Units’, ‘Reporting_Limit’, ‘Validation_Level’, ‘Result_Final_Txt’, ‘Result_Qualifier_Final’, ‘RL_Comparison’].

Our analysis is focussed on the ‘Results_Final_Txt’ which represents the concentration level of the hazardous chemical. The ‘Analyte’ and ‘CAS_NO” (CAS Registry Number) ^[4]^ help identify the chemicals. There are 134 unique chemicals. The records before 10/02/2023 are used for the training set(1166) and the remaining are used for the validation set(656).

### EPA Lab Results: Soil and Sediment (through 02/10/23) - East Palestine, OH Response (csv) ^[3]^

(https://www.epa.gov/oh/soil-and-sediment-sampling-data-east-palestine-ohio-train-derailment) The dataset contains 1082 records starting from 08/02/2023 to 10/02/2023. The dataset contains following features: [‘Location’, ‘Samp_No’, ‘SampleDate_txt’, ‘SampleTime’, ‘Matrix’, ‘SampleMedia’, ‘Activity’, Analytical_Method’, ‘CAS_NO’, ‘Analyte’, ‘Result_Units’, ‘Reporting_Limit’, ‘Validation_Level’, ‘Result_Final_Txt’, ‘Result_Qualifier_Final’, ‘RL_Comparison’].

Our analysis is focussed on the ‘Results_Final_Txt’ which represents the concentration level of the hazardous chemical. The ‘Analyte’ and ‘CAS_NO” (CAS Registry Number) ^[4]^ help identify the chemicals. There are 128 unique chemicals. The records before 10/02/2023 are used for the training set(512) and the remaining are used for the validation set(570).

We combined the ‘SampleDate_txt’ and ‘SampleTime’ features into a single feature called ‘Date-Time’

### Model & Results

#### Air Sample

We use the ‘Analyte’ feature to group the records for each chemical. There are 49 unique hazardous chemicals. We trained separate models for every chemical. We selected the following features for the prediction of our target variable ‘Result_Final_Txt’. ‘Location’, ‘Date_Time’ ‘SampleMedia’, ‘Analytical_Method’, ‘Reporting_Limit’, ‘Validation_Level’, ‘, ‘RL_Comparison’. We use the Augmented Dickey–Fuller test whether the time series is stationary or not. ^[5]^ After experimentation with several forecasting models, ARIMA, SARIMA, LSTM Forecasting ^[8]^. We finally selected the FBProphet forecasting model as it best fitted our training data.FBProphet ^[6][7]^ is a time series forecasting model developed by Facebook’s Data Science team. It is designed to make accurate predictions of future values based on historical time series data, while also handling missing values, outliers, and trend changes. FBProphet uses a decomposable time series model with three main components: trend, seasonality, and holidays. It employs Bayesian inference to fit the model and provides uncertainty intervals for the predictions. We also use the metrics mean square error and mean absolute percentage error ^[9]^ to evaluate the performance of the model in predicting the target variable ‘Result_Final_Txt’, which tells the concentration level of the hazardous chemical in the air. Fig.1, Fig.2, Fig.3, Fig.4 show the prediction of our model against the test data for 4 different chemicals.

**Fig. 1.**
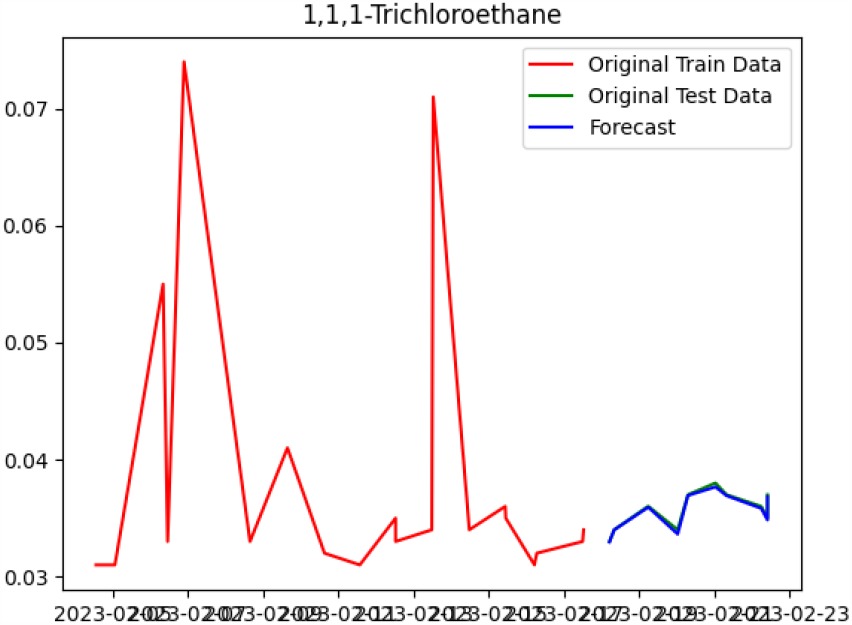

**Fig. 2.**
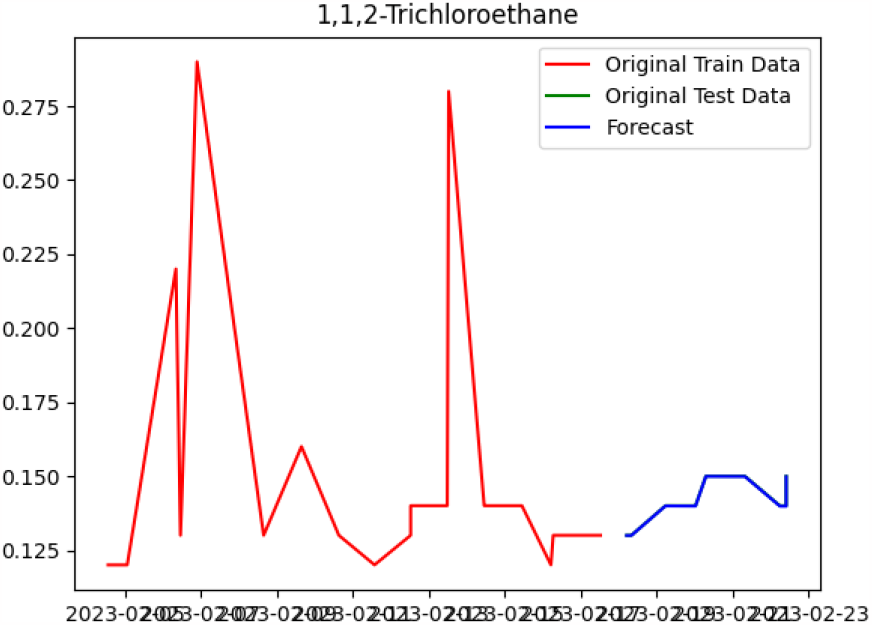

**Fig. 3.**
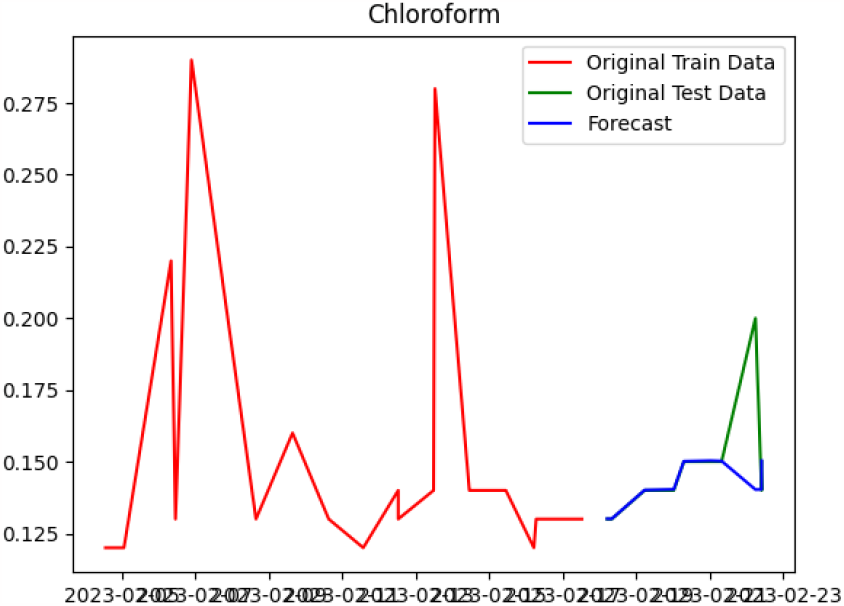

**Fig. 4.**
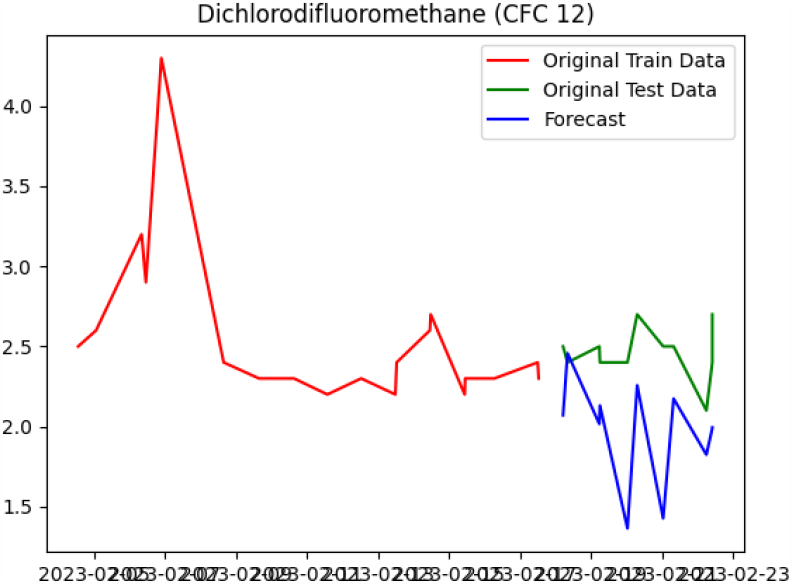

Mean Square Error (MSE) is a commonly used statistical metric that measures the average squared difference between the predicted values and the actual values.

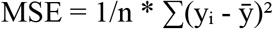

Mean Absolute Percentage Error (MAPE) is a statistical metric that measures the average absolute percentage difference between the predicted values and the actual values.

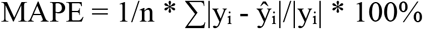

Table 1 shows the performance metrics for each chemical in the Air Sample Data.

**Table 1.**

|  | Chemical | MSE(Air Data) | MAPE(Air Data) |
| --- | --- | --- | --- |
| 0 | Ethylbenzene | 191.0074 | 32.525 |
| 1 | trans-1,2-Dichloroethene | 0.0 | 0.004 |
| 2 | 1,1-Dichloroethane | 0.0 | 0.004 |
| 3 | Methyl tert-butyl ether | 0.0 | 0.004 |
| 4 | cis-1,2-Dichloroethene | 0.0 | 0.004 |
| 5 | Chloroform | 0.0003 | 0.0286 |
| 6 | Tetrachloroethene | 0.1453 | 4.604 |
| 7 | Chlorobenzene | 0.0 | 0.0008 |
| 8 | 1,3,5-Trimethylbenzene | 0.0006 | 0.1302 |
| 9 | Styrene | 0.0102 | 0.4133 |
| 10 | o-Xylene | 73.4363 | 17.6687 |
| 11 | 1,1,2,2-Tetrachloroethane | 0.0 | 0.004 |
| 12 | 1,1,2-Trichlorotrifluoroethane | 0.0055 | 0.1287 |
| 13 | 1,2,4-Trimethylbenzene | 0.0705 | 1.2503 |
| 14 | 1,3-Dichlorobenzene | 0.0 | 0.001 |
| 15 | 1,4-Dichlorobenzene | 2.2509 | 8.1583 |
| 16 | 1,2-Dichlorobenzene | 0.0 | 0.004 |
| 17 | 1,2-Dibromo 3-Chloropropane | 0.0 | 0.0016 |
| 18 | 1,2,4-Trichlorobenzene | 0.0 | 0.0017 |
| 19 | m,p-Xylenes | 2821.5551 | 35.8002 |
| 20 | Methylene chloride | 135.6987 | 26.3074 |
| 21 | cis-1,3-Dichloropropene | 0.0 | 0.0001 |
| 22 | Dibromochloromethane | 0.0 | 0.001 |
| 23 | Dichlorodifluoromethane (CFC 12) | 0.3424 | 0.2019 |
| 24 | Chloromethane | 0.0352 | 0.4056 |
| 25 | Vinyl Chloride | 26.423 | 49.5516 |
| 26 | 1,3-Butadiene | 0.057 | 2.0566 |
| 27 | Bromomethane | 0.0005 | 0.4346 |
| 28 | Chloroethane | 0.0002 | 0.2781 |
| 29 | Acrolein | 0.2012 | 0.8925 |
| 30 | Acetone | 6.7423 | 0.64 |
| 31 | Trichlorofluoromethane | 56.5654 | 4.0665 |
| 32 | 1,2-Dibromoethane | 0.0 | 0.001 |
| 33 | 1,1-Dichloroethene | 0.0 | 0.001 |
| 34 | Benzene | 16.8874 | 3.1617 |
| 35 | Carbon Tetrachloride | 0.0106 | 0.198 |
| 36 | 1,2-Dichloropropane | 0.0294 | 0.2135 |
| 37 | Bromodichloromethane | 0.0 | 0.001 |
| 38 | Trichloroethene | 0.0208 | 2.7841 |
| 39 | 1,4-Dioxane | 0.0 | 0.0032 |
| 40 | trans-1,3-Dichloropropene | 0.0 | 0.0001 |
| 41 | 1,1,2-Trichloroethane | 0.0 | 0.0008 |
| 42 | Toluene | 121.2752 | 1.6103 |
| 43 | 1,2-Dichloroethane | 0.0911 | 2.2759 |
| 44 | 1,1,1-Trichloroethane | 0.0 | 0.004 |
| 45 | Hexachlorobutadiene | 0.0 | 0.0016 |
| 46 | Naphthalene | 0.6466 | 3.1154 |

#### Water Sample

We use the exact same procedure for the Water Sample Data. We use the ‘Analyte’ feature to group the records for each chemical. There are 134 unique hazardous chemicals. We trained separate models for every chemical.Fig.5, Fig.6, Fig.7, Fig.8 show the prediction of our model against the test data for 4 different chemicals.

**Fig. 5.**
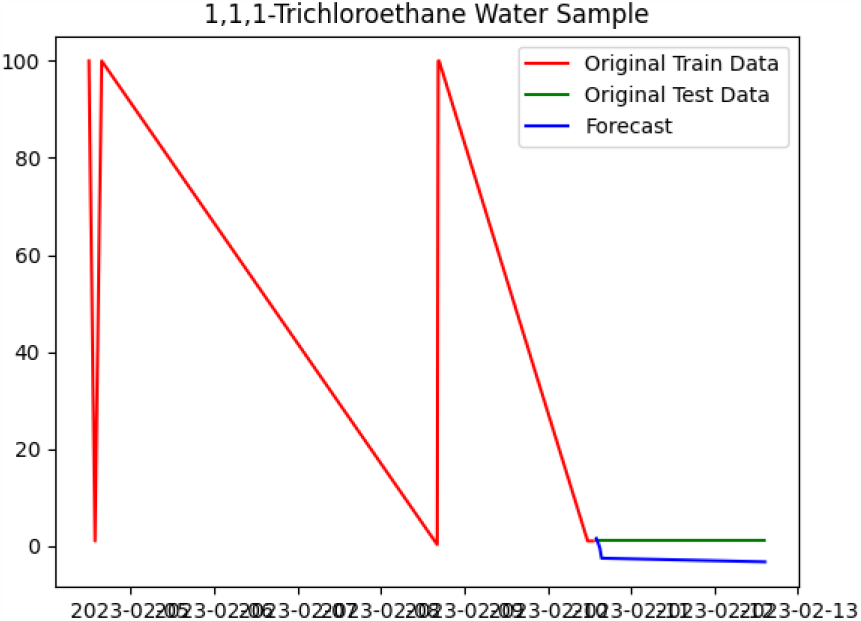

**Fig. 6.**
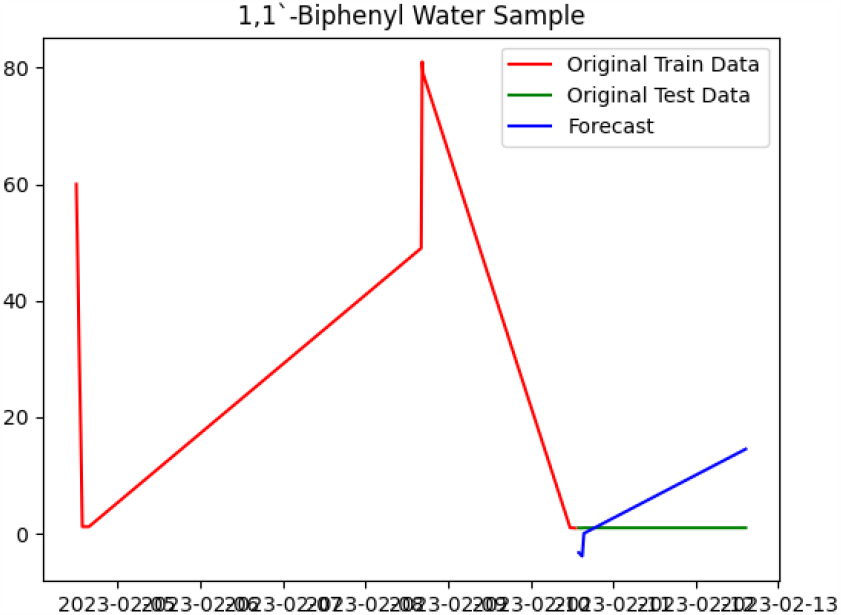

**Fig. 7.**
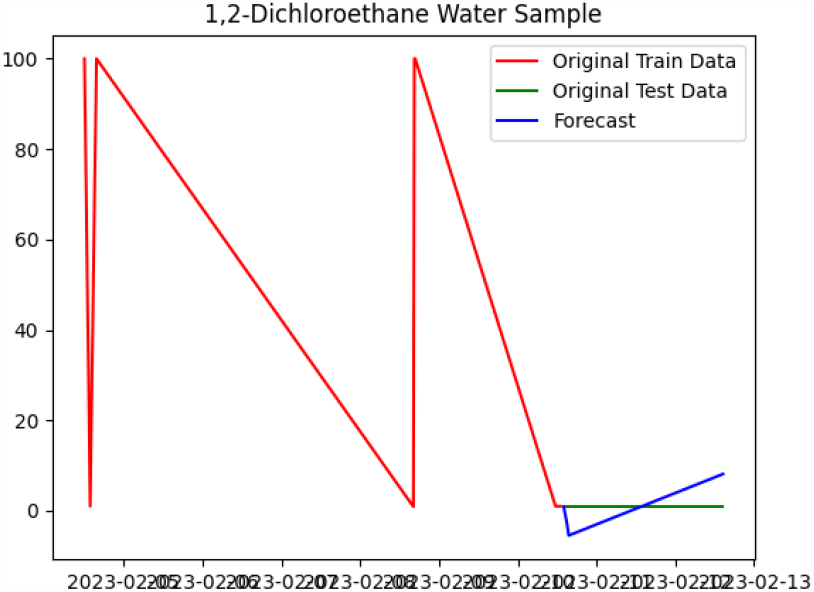

**Fig. 8.**
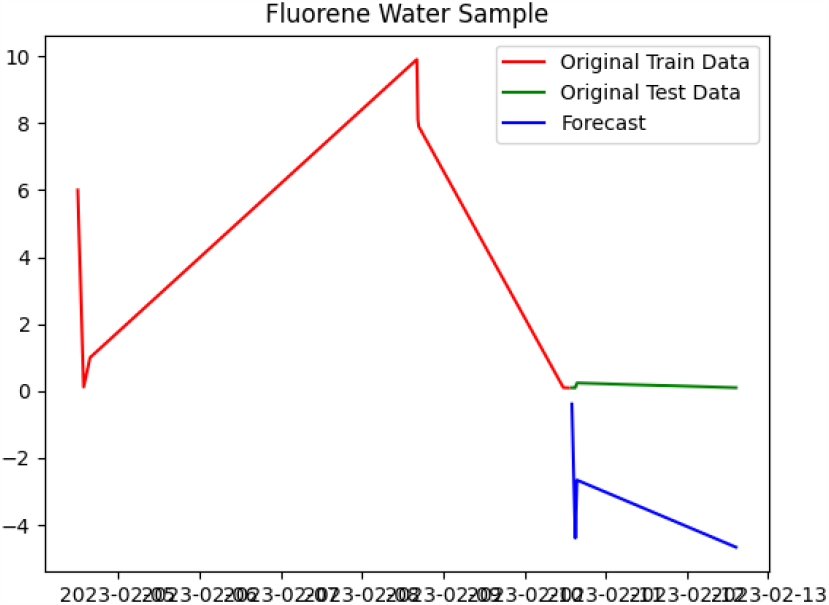

Table 2, 3, 4, 5 shows the performance metrics for each chemical in the Water Sample Data.

**Table 2.**

|  | Chemical | MSE(Water Data) | MAPE(Water Data) |
| --- | --- | --- | --- |
| 0 | 1,2-Dichloroethane | 24.66300000 | 4.3203 |
| 1 | Bromoform | 7.03750000 | 2.238 |
| 2 | 1,4-Dichlorobenzene | 7.03750000 | 2.238 |
| 3 | 1,3-Dichlorobenzene | 7.03750000 | 2.238 |
| 4 | 1,3,5-Trimethylbenzene | 24.66300000 | 4.3203 |
| 5 | 1,2-Dichloropropane | 7.03750000 | 2.238 |
| 6 | 1,2-Dichlorobenzene | 7.03750000 | 2.238 |
| 7 | 1,2-Dibromoethane | 7.03750000 | 2.238 |
| 8 | 2-Nitroaniline | 49.50020000 | 5.7653 |
| 9 | 2-Methylphenol | 49.50020000 | 5.7653 |
| 10 | 2-Methylnaphthalene | 68.87890000 | 74.8085 |
| 11 | 2-Chlorophenol | 49.50020000 | 5.7653 |
| 12 | 2-Chloronaphthalene | 0.91470000 | 9.5397 |
| 13 | 2,6-Dinitrotoluene | 49.50020000 | 5.7653 |
| 14 | 2,4-Dinitrotoluene | 49.50020000 | 5.7653 |
| 15 | 1-Methylnaphthalene | 2.58340000 | 14.0021 |
| 16 | 1,4-Dioxane | 380.15200000 | 3.4628 |
| 17 | 1,2,4,5-Tetrachlorobenzene | 485.38650000 | 3.9184 |
| 18 | Pyridine | 202.56250000 | 1.4139 |
| 19 | 1,1,1-Trichloroethane | 7.03750000 | 2.238 |
| 20 | 1,1,2,2-Tetrachloroethane | 7.03750000 | 2.238 |
| 21 | 1,1,2-Trichloroethane | 7.03750000 | 2.238 |
| 22 | 1,1,2-Trichlorotrifluoroethane | 7.03750000 | 2.238 |
| 23 | 1,1-Dichloroethane | 7.03750000 | 2.238 |
| 24 | Bromomethane | 24.66300000 | 4.3203 |
| 25 | 1,1-Dichloroethene | 7.03750000 | 2.238 |
| 26 | 1,2,3-Trichlorobenzene | 24.66300000 | 4.3203 |
| 27 | 1,2,4-Trichlorobenzene | 24.66300000 | 4.3203 |
| 28 | 1,2,4-Trimethylbenzene | 7.03750000 | 2.238 |
| 29 | 1,2-Dibromo 3-Chloropropane | 24.66300000 | 4.3203 |
| 30 | GRO (C6-C10) | 33870318770.65290070 | 288.7863 |

**Table 3.**

|  |  |  |  |
| --- | --- | --- | --- |
| 31 | 1,1'-Biphenyl | 49.50020000 | 5.7653 |
| 32 | 1,1-Dichloropropene | 58.70210000 | 5.8189 |
| 33 | Carbon disulfide | 7.03750000 | 2.238 |
| 34 | Carbon Tetrachloride | 7.03750000 | 2.238 |
| 35 | Chlorobenzene | 7.03750000 | 2.238 |
| 36 | 2-Butanone | 4.36820000 | 0.4098 |
| 37 | 2-Hexanone | 0.81900000 | 0.1531 |
| 38 | 4-Methyl-2-pentanone | 7.03750000 | 2.238 |
| 39 | Acetone | 921.81860000 | 2.3764 |
| 40 | Benzene | 7.03750000 | 2.238 |
| 41 | Bromochloromethane | 7.03750000 | 2.238 |
| 42 | Xylenes, Total | 5.97870000 | 1.1224 |
| 43 | Bromodichloromethane | 7.03750000 | 2.238 |
| 44 | m,p-Xylenes | 0.89520000 | 0.3526 |
| 45 | Methyl acetate | 9.26210000 | 1.1911 |
| 46 | Methyl tert-butyl ether | 7.03750000 | 2.238 |
| 47 | Methylcyclohexane | 7.03750000 | 2.238 |
| 48 | Methylene chloride | 115332.82640000 | 51.2957 |
| 49 | o-Xylene | 7.03750000 | 2.238 |
| 50 | Isopropylbenzene | 7.03750000 | 2.238 |
| 51 | Pyrene | 29.66720000 | 48.1067 |
| 52 | Vinyl Chloride | 10895673.57030000 | 1897.9536 |
| 53 | Trichloroethene | 7.03750000 | 2.238 |
| 54 | Chloroethane | 24.66300000 | 4.3203 |
| 55 | Chloroform | 7.03750000 | 2.238 |
| 56 | Chloromethane | 24.66300000 | 4.3203 |
| 57 | cis-1,2-Dichloroethene | 7.03750000 | 2.238 |
| 58 | cis-1,3-Dichloropropene | 7.03750000 | 2.238 |
| 59 | Cyclohexane | 4.42390000 | 0.8032 |
| 60 | Trichlorofluoromethane | 7.03750000 | 2.238 |
| 61 | Dibromochloromethane | 7.03750000 | 2.238 |
| 62 | Ethylbenzene | 7.03750000 | 2.238 |
| 63 | Styrene | 7.03750000 | 2.238 |

**Table 4.**

|  |  |  |  |
| --- | --- | --- | --- |
| 64 | Tetrachloroethene | 7.03750000 | 2.238 |
| 65 | Toluene | 7.03750000 | 2.238 |
| 66 | trans-1,2-Dichloroethene | 7.03750000 | 2.238 |
| 67 | trans-1,3-Dichloropropene | 7.03750000 | 2.238 |
| 68 | Dibromodifluoromethane | 58.70210000 | 5.8189 |
| 69 | Phenol | 49.50020000 | 5.7653 |
| 70 | 1,2,3-Trichloropropane | 7.03750000 | 2.238 |
| 71 | Pentachlorophenol | 485.38650000 | 3.9184 |
| 72 | Caprolactam | 485.38650000 | 3.9184 |
| 73 | Butyl benzyl phthalate | 49.50020000 | 5.7653 |
| 74 | Bis(2-ethylhexyl)phthalate | 108.27980000 | 4.4252 |
| 75 | Bis(2-chloroethyl)ether | 49.50020000 | 5.7653 |
| 76 | Bis(2-chloroethoxy)methane | 49.50020000 | 5.7653 |
| 77 | Benzo(k)fluoranthene | 0.46910000 | 6.9082 |
| 78 | Benzo(g,h,i)perylene | 2.60490000 | 16.0966 |
| 79 | Benzo(b)fluoranthene | 20.02340000 | 36.5454 |
| 80 | Benzo(a)pyrene | 0.61990000 | 7.198 |
| 81 | Benzo(a)anthracene | 137.37150000 | 111.9364 |
| 82 | Benzaldehyde | 49.50020000 | 5.7653 |
| 83 | Atrazine | 49.50020000 | 5.7653 |
| 84 | Anthracene | 49.51000000 | 64.9035 |
| 85 | Phenanthrene | 820.79500000 | 172.379 |
| 86 | Acenaphthylene | 14.14230000 | 32.1705 |
| 87 | Acenaphthene | 0.82810000 | 7.9685 |
| 88 | 4-Nitrophenol | 485.38650000 | 3.9184 |
| 89 | 4-Nitroaniline | 49.50020000 | 5.7653 |
| 90 | 4-Chlorophenyl phenyl ether | 49.50020000 | 5.7653 |
| 91 | 4-Chloroaniline | 28.48450000 | 4.9227 |
| 92 | 4-Chloro-3-methylphenol | 49.50020000 | 5.7653 |
| 93 | 4-Bromophenyl phenyl ether | 49.50020000 | 5.7653 |
| 94 | 4,6-Dinitro-2-methylphenol | 49.50020000 | 5.7653 |
| 95 | 3-Nitroaniline | 49.50020000 | 5.7653 |
| 96 | 3,3'-Dichlorobenzidine | 485.38650000 | 3.9184 |

**Table 5.**

|  |  |  |  |
| --- | --- | --- | --- |
| 97 | 3&4-Methylphenol | 49.50020000 | 5.7653 |
| 98 | 2-Nitrophenol | 49.50020000 | 5.7653 |
| 99 | Carbazole | 49.50020000 | 5.7653 |
| 100 | Chrysene | 1.81720000 | 12.239 |
| 101 | Acetophenone | 49.50020000 | 5.7653 |
| 102 | Hexachloroethane | 49.50020000 | 5.7653 |
| 103 | Dibenzo(a,h)anthracene | 0.91470000 | 9.5397 |
| 104 | N-Nitrosodiphenylamine | 49.50020000 | 5.7653 |
| 105 | N-Nitrosodi-n-propylamine | 49.50020000 | 5.7653 |
| 106 | Nitrobenzene | 49.50020000 | 5.7653 |
| 107 | Naphthalene | 4.17720000 | 10.2575 |
| 108 | Isophorone | 380.15200000 | 3.4628 |
| 109 | Indeno(1,2,3-cd)pyrene | 4.31440000 | 18.5967 |
| 110 | Hexachlorocyclopentadiene | 380.15200000 | 3.4628 |
| 111 | Hexachlorobutadiene | 49.50020000 | 5.7653 |
| 112 | Hexachlorobenzene | 49.50020000 | 5.7653 |
| 113 | Fluoranthene | 224.06530000 | 112.7274 |
| 114 | 2,4-Dinitrophenol | 380.15200000 | 3.4628 |
| 115 | Fluorene | 14.19970000 | 31.5878 |
| 116 | 2,4-Dichlorophenol | 49.50020000 | 5.7653 |
| 117 | Dibenzofuran | 49.50020000 | 5.7653 |
| 118 | Diethyl phthalate | 49.50020000 | 5.7653 |
| 119 | Dimethyl phthalate | 49.50020000 | 5.7653 |
| 120 | Di-n-butyl phthalate | 39.39040000 | 5.9788 |
| 121 | Di-n-octyl phthalate | 49.50020000 | 5.7653 |
| 122 | 2,4-Dimethylphenol | 49.50020000 | 5.7653 |
| 123 | ORO (C28-C40) | 42558591339820.28906250 | 20595.075 |
| 124 | DRO (C10-C28) | 78432168272756.54687500 | 36134.811 |
| 125 | 2,2`-Oxybis(1-chloropropane) | 49.50020000 | 5.7653 |
| 126 | 2,3,4,6-Tetrachlorophenol | 49.50020000 | 5.7653 |
| 127 | 2,4,5-Trichlorophenol | 49.50020000 | 5.7653 |
| 128 | 2,4,6-Trichlorophenol | 49.50020000 | 5.7653 |

#### Soil Sample

We use the exact same procedure for the Soil Sample Data. We use the ‘Analyte’ feature to group the records for each chemical. There are 128 unique hazardous chemicals. We trained separate models for every chemical.Fig.9, Fig.10, Fig.11, Fig.12 show the prediction of our model against the test data for 4 different chemicals.

**Fig. 9.**
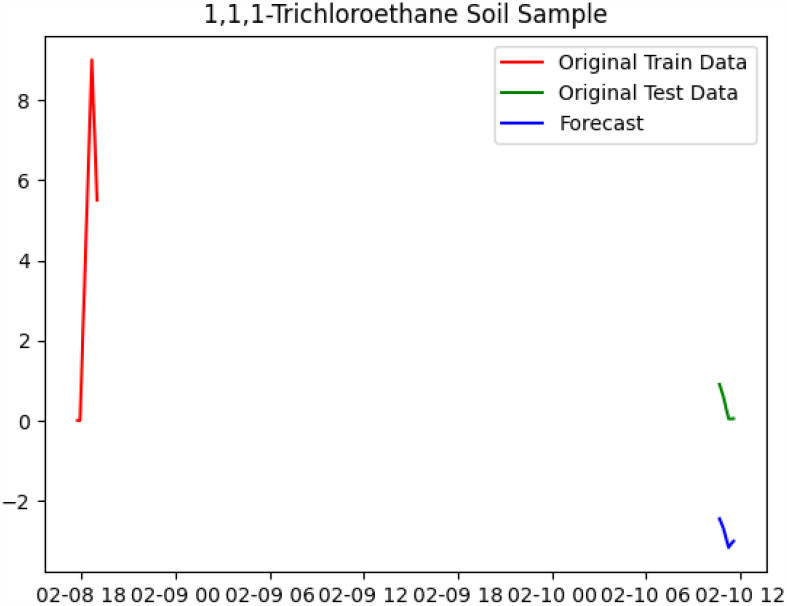

**Fig. 10.**
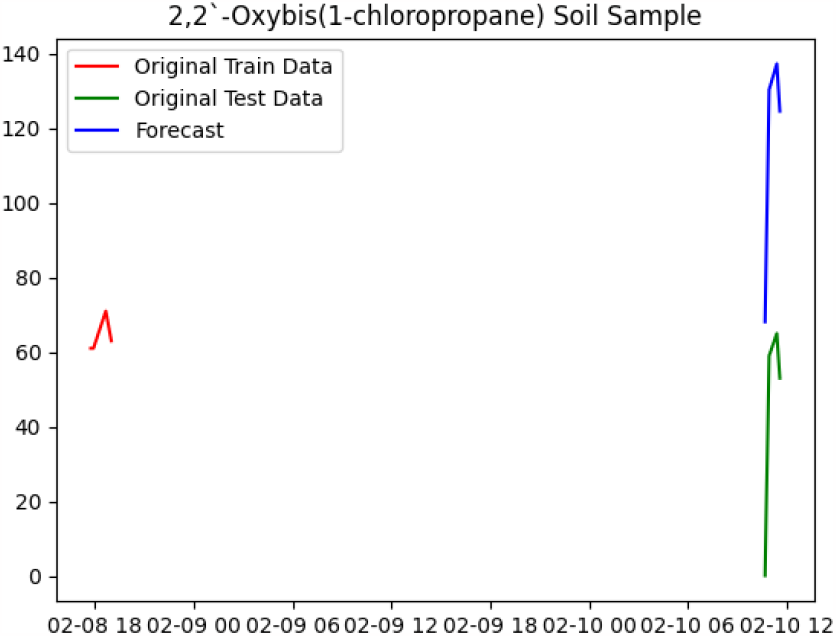

**Fig. 11.**
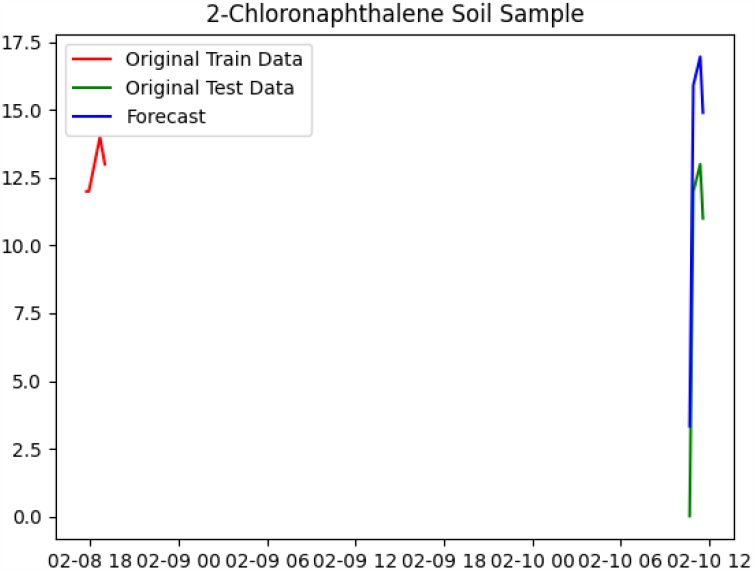

**Fig. 12.**
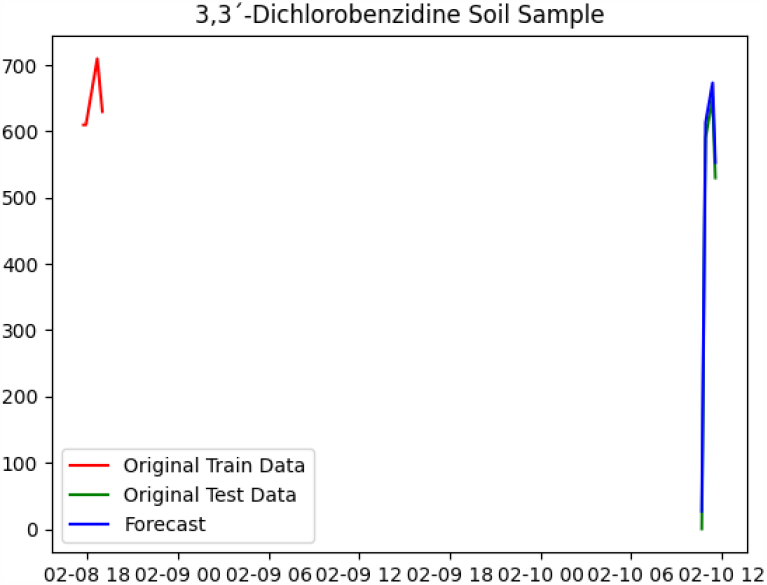

Table 6, 7, 8, 9 shows the performance metrics for each chemical in the Soil Sample Data.

**Table 6.**

|  | Chemical | MSE(Soil Data) | MAPE(Soil Data) |
| --- | --- | --- | --- |
| 0 | 2-Butanone | 43.2635 | 13.2777 |
| 1 | cis-1,2-Dichloroethene | 10.2881 | 40.352 |
| 2 | Chloromethane | 0.0114 | 0.4255 |
| 3 | Bromomethane | 0.0114 | 0.4255 |
| 4 | Fluoranthene | 2621.26 | 101.3533 |
| 5 | Hexachlorobutadiene | 5021.2002 | 387.6414 |
| 6 | Hexachlorobenzene | 5021.2002 | 387.6414 |
| 7 | Dichlorodifluoromethane (CFC 12) | 0.0114 | 0.4255 |
| 8 | Hexachlorocyclopentadiene | 5021.2002 | 387.6414 |
| 9 | Hexachloroethane | 5021.2002 | 387.6414 |
| 10 | Fluorene | 14.3053 | 29.7113 |
| 11 | Ethylbenzene | 10.2881 | 40.352 |
| 12 | Methylcyclohexane | 0.0107 | 0.7653 |
| 13 | Methyl tert-butyl ether | 10.2881 | 40.352 |
| 14 | Indeno(1,2,3-cd)pyrene | 5018.5557 | 203.2561 |
| 15 | Methylene chloride | 10.5566 | 4.9879 |
| 16 | o-Xylene | 1.3968 | 12.7102 |
| 17 | Styrene | 10.2881 | 40.352 |
| 18 | trans-1,3-Dichloropropene | 10.2881 | 40.352 |
| 19 | Trichloroethene | 10.2881 | 40.352 |
| 20 | m,p-Xylenes | 0.0 | 0.0142 |
| 21 | Methyl acetate | 10.5566 | 4.9879 |
| 22 | Tetrachloroethene | 10.2881 | 40.352 |
| 23 | Isopropylbenzene | 10.2881 | 40.352 |
| 24 | Isophorone | 5021.6528 | 78.2781 |
| 25 | Naphthalene | 14.2888 | 21.8916 |
| 26 | Nitrobenzene | 5021.6528 | 78.2781 |
| 27 | Trichlorofluoromethane | 10.2881 | 40.352 |
| 28 | Vinyl Chloride | 128895.3877 | 1488.9461 |
| 29 | Xylenes, Total | 75.8403 | 33.6355 |
| 30 | 1,4-Dichlorobenzene | 10.2881 | 40.352 |

**Table 7.**
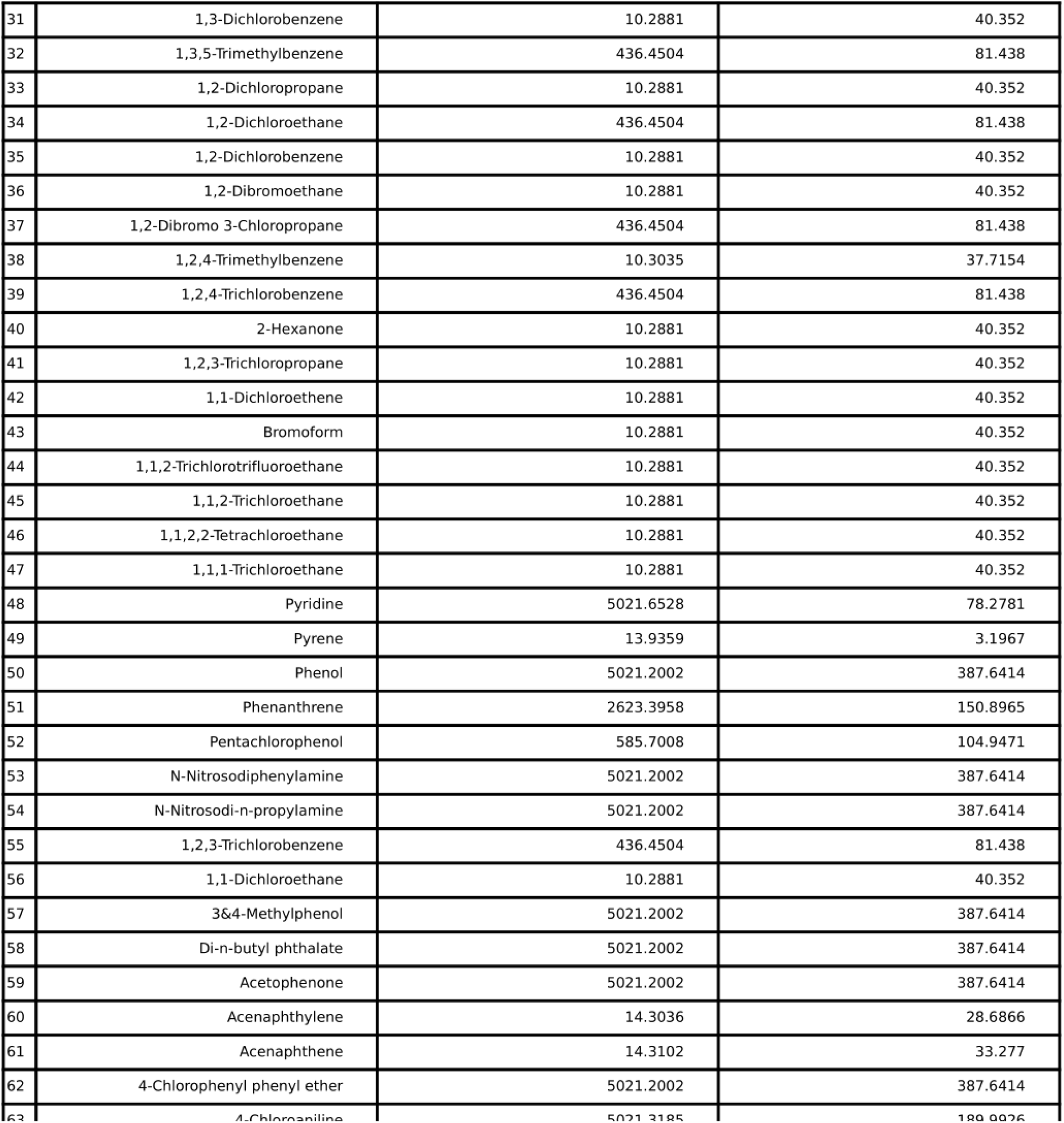

**Table 8.**
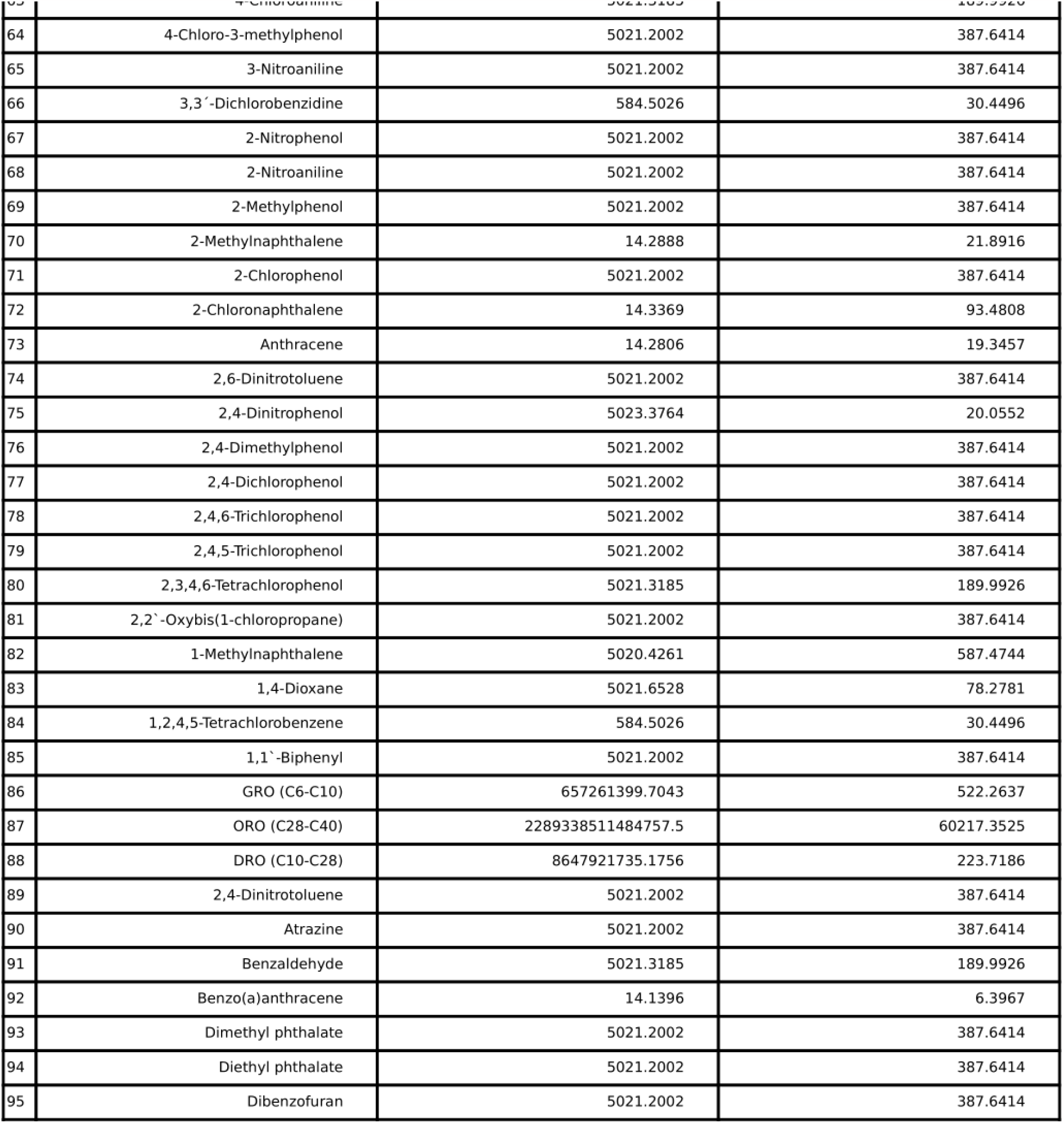

**Table 9.**

|  |  |  |  |
| --- | --- | --- | --- |
| 96 | Butyl benzyl phthalate | 5021.3185 | 189.9926 |
| 97 | Bis(2-ethylhexyl)phthalate | 5021.2002 | 387.6414 |
| 98 | Bis(2-chloroethyl)ether | 5021.2002 | 387.6414 |
| 99 | Bis(2-chloroethoxy)methane | 5021.2002 | 387.6414 |
| 100 | Benzo(k)fluoranthene | 14.2381 | 12.0548 |
| 101 | Benzo(g,h,i)perylene | 14.2397 | 12.2321 |
| 102 | Benzo(b)fluoranthene | 14.0446 | 4.3756 |
| 103 | Benzo(a)pyrene | 14.1717 | 7.5604 |
| 104 | Dibromochloromethane | 10.2881 | 40.352 |
| 105 | Cyclohexane | 0.0114 | 0.4255 |
| 106 | cis-1,3-Dichloropropene | 10.2881 | 40.352 |
| 107 | Chloroform | 10.2881 | 40.352 |
| 108 | Chloroethane | 436.4504 | 81.438 |
| 109 | Chlorobenzene | 10.2881 | 40.352 |
| 110 | 4,6-Dinitro-2-methylphenol | 5021.2002 | 387.6414 |
| 111 | 4-Bromophenyl phenyl ether | 5021.2002 | 387.6414 |
| 112 | 4-Nitroaniline | 5021.6528 | 78.2781 |
| 113 | 4-Nitrophenol | 584.5026 | 30.4496 |
| 114 | Carbazole | 5021.2002 | 387.6414 |
| 115 | Chrysene | 14.1556 | 6.9301 |
| 116 | Di-n-octyl phthalate | 5021.2002 | 387.6414 |
| 117 | Dibenzo(a,h)anthracene | 14.3136 | 36.171 |
| 118 | Acetone | 58181.8616 | 550.6706 |
| 119 | Benzene | 2610992.6339 | 3135.9938 |
| 120 | Bromochloromethane | 10.2881 | 40.352 |
| 121 | Bromodichloromethane | 10.2881 | 40.352 |
| 122 | Carbon disulfide | 10.2881 | 40.352 |
| 123 | Carbon Tetrachloride | 10.2881 | 40.352 |
| 124 | 4-Methyl-2-pentanone | 10.2881 | 40.352 |
| 125 | Caprolactam | 584.4983 | 74.3885 |
| 126 | trans-1,2-Dichloroethene | 10.2881 | 40.352 |
| 127 | Toluene | 10.3493 | 32.64 |

## Air Sample with Water Sample

Here we try to predict the target for Air Sample data by combining both Air Sample and Water Sample Data and using Water Sample data as an auxiliary variable. Table 10, 11 shows the performance metrics for each chemical

**Table 10.**

|  | Chemical | MSE(Air Data) | MAPE(Air Data) | MSE(Air & Water Data) | MAPE(Air & WaterData) |
| --- | --- | --- | --- | --- | --- |
| 0 | 1,1-Dichloroethene | 0.0 | 0.0132 | 0.0 | 0.0023 |
| 1 | 1,2-Dibromoethane | 0.0 | 0.004 | 0.0 | 0.0023 |
| 2 | Trichlorofluoromethane | 53.0247 | 139.8308 | 101.4218 | 5.7626 |
| 3 | Chloroethane | 1.3321 | 0.9742 | 0.002 | 0.8072 |
| 4 | Bromomethane | 0.0004 | 0.4147 | 0.0006 | 0.5289 |
| 5 | Benzene | 11.369 | 64.9293 | 214.1085 | 11.8429 |
| 6 | Acetone | 29.4171 | 6.9373 | 20.6929 | 0.9085 |
| 7 | Carbon Tetrachloride | 13.6164 | 0.9098 | 0.0178 | 0.2566 |
| 8 | 1,1,2-Trichloroethane | 0.0886 | 0.6756 | 0.0 | 0.0052 |
| 9 | Bromodichloromethane | 0.0113 | 0.7481 | 0.0 | 0.0148 |
| 10 | Trichloroethene | 0.0224 | 3.221 | 0.0228 | 3.1453 |
| 11 | 1,4-Dioxane | 0.0104 | 2.6689 | 0.0 | 0.01 |
| 12 | trans-1,3-Dichloropropene | 0.005 | 0.4983 | 0.0 | 0.0028 |
| 13 | Toluene | 9.7706 | 38.2197 | 137.3863 | 1.9817 |
| 14 | 1,2-Dichloroethane | 129.019 | 0.8709 | 0.1358 | 2.8332 |
| 15 | 1,1,1-Trichloroethane | 0.0011 | 0.4695 | 0.0 | 0.0023 |
| 16 | Hexachlorobutadiene | 0.0113 | 2.9658 | 0.0 | 0.0021 |
| 17 | Naphthalene | 0.6483 | 3.193 | 0.0758 | 1.5787 |
| 18 | Vinyl Chloride | 10.9528 | 17.5858 | 1160.488 | 274.6528 |
| 19 | 1,2-Dichloropropane | 13.2683 | 0.6304 | 0.0296 | 0.2252 |
| 20 | Chloromethane | 0.1201 | 7.4631 | 0.1244 | 0.8034 |
| 21 | 1,2-Dibromo 3-Chloropropane | 0.0499 | 0.5887 | 0.0 | 0.0017 |

**Table 11.**

|  |  |  |  |  |  |
| --- | --- | --- | --- | --- | --- |
| 22 | Dibromochloromethane | 0.0113 | 0.7489 | 0.0 | 0.0023 |
| 23 | Dichlorodifluoromethane (CFC 12) | 3.8515 | 54.3666 | 0.3409 | 0.2017 |
| 24 | trans-1,2-Dichloroethene | 5.9189 | 0.9855 | 0.0 | 0.0148 |
| 25 | 1,1-Dichloroethane | 0.0 | 0.001 | 0.0 | 0.0148 |
| 26 | Methyl tert-butyl ether | 0.0 | 0.001 | 0.0 | 0.0148 |
| 27 | cis-1,2-Dichloroethene | 0.0 | 0.004 | 0.0 | 0.0023 |
| 28 | Chloroform | 0.0113 | 2.9658 | 0.0003 | 0.0325 |
| 29 | Tetrachloroethene | 0.1308 | 2.1817 | 0.2734 | 4.3371 |
| 30 | Chlorobenzene | 0.0221 | 1.2708 | 0.0 | 0.0068 |
| 31 | 1,3,5-Trimethylbenzene | 0.0008 | 0.1481 | 0.0014 | 0.1722 |
| 32 | Styrene | 0.0139 | 0.5708 | 0.0096 | 0.4231 |
| 33 | o-Xylene | 74.9303 | 29.5312 | 127.19 | 21.2755 |
| 34 | Ethylbenzene | 187.6134 | 22.8647 | 195.2935 | 32.9108 |
| 35 | 1,1,2-Trichlorotrifluoroethane | 0.0731 | 1.5922 | 0.0069 | 0.1471 |
| 36 | 1,2,4-Trimethylbenzene | 0.1045 | 0.5228 | 0.2369 | 1.3977 |
| 37 | 1,3-Dichlorobenzene | 0.058 | 0.8068 | 0.0 | 0.0148 |
| 38 | 1,4-Dichlorobenzene | 2.5723 | 32.0237 | 3.645 | 10.152 |
| 39 | 1,2-Dichlorobenzene | 0.0782 | 0.7549 | 0.0 | 0.0023 |
| 40 | 1,2,4-Trichlorobenzene | 0.0013 | 0.9929 | 0.0 | 0.0091 |
| 41 | m,p-Xylenes | 2781.5664 | 347.5855 | 9553.0109 | 60.689 |
| 42 | Methylene chloride | 138.9483 | 16.166 | 505.7796 | 50.0844 |
| 43 | 1,1,2,2-Tetrachloroethane | 0.1778 | 0.898 | 0.0 | 0.0148 |
| 44 | cis-1,3-Dichloropropene | 0.0013 | 0.9904 | 0.0 | 0.0028 |

The mean and median for MSE for Air Sample Data is 73.4789 and 0.0006 respectively. The mean and median for MAPE for Air Sample Data is 4.2328 and 0.198 respectively.

The mean and median for MSE for Water Sample Data is 938175511251.79 and 28.4845 respectively.

The mean and median for MAPE for Water Sample Data is 466.434 and 5.7653 respectively.

The mean and median for MSE for Soil Sample Data is17885529841550.645 and 436.4504 respectively.

The mean and median for MAPE for Soil Sample Data is 663.1171 and 57.37025 respectively.

The mean and median for MSE for combined Air and Water Ssmple Data is 267.11765 and 0.0014 respectively.

The mean and median for MAPE for Soil Sample Data is 10.8287 and 0.1722 respectively.

## Conclusion

In conclusion the time series forecasting of Ohio train derailment data has provided valuable insights into the patterns of the hazardous chemical present in air, water and soil.

The time series forecasting models used in this research have demonstrated strong predictive power when using Air Sample Data and Air Sample Data with Water Sample Data used as an auxiliary variable.

The relatively poor performance of the soil and water model with respect to the air model can be alluded to the fact that there is a lack of temporal data for both soil and water. While Air Sample Data has a total data collection of 18 days, data samples for water and soil have only been collected for 11 and 2 days respectively.

We hope that this research comes to use and the models can be used for forecasting hazardous chemical concentration over time.

## Data Availability

All data produced are available online on EPA website.

